# Young and invisible: an explanatory model for service engagement by people who inject drugs in India

**DOI:** 10.1101/2021.02.23.21252222

**Authors:** Lakshmi Ganapathi, Aylur K Srikrishnan, Clarissa Martinez, Gregory M Lucas, Shruti H Mehta, Vinita Verma, Allison McFall, Kenneth H. Mayer, Areej Hassan, Shobini Rajan, Conall O’Cleirigh, Sion Kim Harris, Sunil S Solomon

**Author notes:** Corresponding author: Lakshmi Ganapathi, Division of Infectious Diseases, Boston Children’s Hospital, 300 Longwood Avenue, Boston, MA 02115.

## Abstract

**Introduction:** The HIV epidemic in India is concentrated in key populations such as people who inject drugs (PWID). New HIV infections are high among young PWID (≤ 30 years of age), who are hard to engage in services. We assessed perspectives of young PWID across three Indian cities representing historic and emerging drug use epidemics to guide development of youth-specific services.

**Methods:** We conducted focus group discussions (FGDs) with PWID (ages 18-35 years) and staff at venues offering services to PWID in three cities (Aizawl and Imphal, Northeast India and Amritsar, Northwest India). A semi-structured interview guide was used to elicit participants’ narratives on injection initiation experiences, motivating factors and barriers to seeking harm-reduction services, service-delivery gaps, and recommendations to promote engagement.

Thematic analysis was used to develop an explanatory model for engagement for each temporal stage across the injection continuum: (a) pre-injection initiation, (b) peri-injection initiation and (c) established injection behavior.

**Results:** 43 PWID (81% male, 19% female) and 10 staff members participated in FGDs. Injection initiation followed non-injection opioid dependence. Lack of services for non-injection opioid dependence was a key gap in the pre-injection initiation phase. Lack of knowledge and reliance on informal sources for injecting equipment were key reasons for non-engagement in the peri-injection phase. Additionally, low risk perception resulted in low motivation to seek services.

Psychosocial and structural factors shaped engagement after established injection. Housing and food insecurity, and stigma disproportionately affected female PWID while lack of confidential adolescent friendly services impeded engagement by adolescent PWID.

**Conclusions:** Development of youth-specific services for young PWID in India will need to address unique vulnerabilities and service gaps along each stage of the injection continuum. Scaling-up of tailored services is needed for young female PWID and adolescents, including interventions that prevent injection initiation and provision of confidential harm-reduction services.

**STRENGTHS AND LIMITATIONS:**

- The findings in this study represent some of the first qualitative data to explore engagement with services, specifically among young PWID in India.
- The study was conducted in multiple cities representing older and emerging injection drug use epidemics. The inclusion of multiple cities adds strength to the findings.
- We did not recruit adolescent PWID due to constraints rendered by laws pertaining to informed consent in India.
- Although all PWID we recruited had initiated injection in adolescence or young adulthood, the preponderance of older PWID in our study limits the conclusions we can draw about the needs of adolescent PWID.

## INTRODUCTION

There are approximately 15.6 million people who inject drugs (PWID) globally, with most living in low-and middle-income countries[1,2]. Although injection initiation often happens in adolescence, there are no global prevalence estimates of adolescent PWID, nor comprehensive HIV and hepatitis C estimates in this vulnerable population. The limited available data in several countries indicate that adolescent and young adult PWID bear a disproportionate burden of new infections[3–6]. Finding, engaging and retaining young PWID in comprehensive HIV prevention and treatment programs is therefore critical to curb the HIV epidemic, particularly in countries like India where the epidemic is concentrated in key populations[7].

India has the largest number of opioid users in the world[8]. More recent estimates from the Ministry of Social Justice and Empowerment, Government of India suggest that there are about 850,000 PWID in India[9]. Among PWID, nearly half are youth under the age of thirty[10]. Injection drug use (IDU) is prevalent and the primary driver of the HIV epidemic in the Northeastern states of India[11,12], given their close proximity to heroin production in Southeast Asian countries. Historically, HIV prevention interventions among PWID have focused on this region. However, recent attention has been drawn to rapidly growing IDU with pharmaceutical opioids and burgeoning HIV and hepatitis C epidemics in states outside the Northeastern region previously considered to have low HIV prevalence[13–15]. In a cross-sectional study of ∼15,000 PWID across 15 Indian cities, we described the tendency for injection initiation to occur in adolescence and significantly greater injection and sexual risk behaviors among young adult PWID (i.e., those between ages 18-24 years) compared to older PWID[16]. Young adult PWID were noted to be driving the HIV epidemic in regions with emerging IDU epidemics[16]. Additionally, young adult PWID had low levels of participation in syringe services (SSP) and opioid substitution therapy (OST; OST is the nomenclature commonly used in India as the vast majority of programs provide sublingual buprenorphine, a partial agonist), even in the Northeastern states where such services are widely available[16] via community-based non-governmental organizations (NGOs), which are supported by the National AIDS Control Organization (NACO) and offer HIV prevention and care services to PWID. There is paucity of data on the availability, accessibility and effectiveness of youth-specific services for young PWID in India.

Since 2013, in partnership with NACO and local NGOs, we have established integrated care centers (ICCs) (that offer outpatient single-venue comprehensive HIV prevention and treatment services to PWID across 12 Indian cities) and the work of evaluating their impact is emerging [17]. The World Health Organization has recently described adolescent-specific considerations in the implementation of health services for key populations[18]. Adaptation of these adolescent-specific considerations for young Indian PWID would require understanding of how their unique vulnerabilities are shaped by local contexts of drug use, cultural norms, laws and policies. In addition, their inputs can guide the types of services most likely to engage them. This study’s objective was to explore perspectives of young PWID and PWID service delivery staff in three Indian cities representing older and emerging IDU epidemics to inform development of services for adolescent and young adult PWID.

## METHODS

### Study settings

Informed by our previous research[13], we chose three sites to conduct FGDs with PWID and staff at NGOs offering services to PWID primarily via the ICCs: (1) Aizawl, in Mizoram state; (2) Imphal, in Manipur state (both in the Northeast region representing sites of historical IDU epidemics); (3) Amritsar, in Punjab state (Northwest region, where the IDU epidemic has been more recent). We chose these cities as our prior research identified low median age of injection initiation, corroborated also by other studies that have described youth susceptibility to IDU in these states[19–22]. Therefore, our recruitment efforts were likely to identify PWID who met study criteria (described below).

While all these states are located along heroin trafficking routes given their proximity to the “Golden triangle” countries (i.e., Thailand, Myanmar and Laos) of heroin production in Southeast Asia (Mizoram and Manipur) and the “Golden crescent” countries (i.e., Pakistan and Afghanistan) in the Northwest (Punjab), they differ in socio-economic characteristics. The Northeast region has been characterized for decades by high unemployment among young people despite high educational levels[23]. In addition, longstanding and ongoing socio-political instability in Mizoram and Manipur shape the daily lives of residents, including healthcare delivery. In contrast, an insurgency that arose in the late 1970s and early 1980s in Punjab has long abated and the state has experienced decline in its relative affluence only recently, due to a stagnating agricultural sector and minimal industrialization[24].

The epidemiology of drug use and HIV also varies in these states. The HIV epidemic among male and female PWID is well characterized in the Northeast. Manipur is known to have the highest prevalence of female PWID in India[25,26]. Aizawl in Mizoram state has the fastest growing HIV epidemic in India currently. Services for female PWID, while less developed than those for men, have been in existence longer in the Northeastern states. In contrast, reports of IDU among women in Punjab have emerged recently[27], and gender-specific services are underdeveloped.

### Study population

We employed purposive sampling using combined criterion and maximum variation strategies[28]. Criteria used to recruit PWID were: (1) age between 18 to 24 years, a criterion liberalized to include participants up to 30 years for men and 35 years for women after initial recruitment efforts over the course of a month due to challenges recruiting young adults; (2) IDU initiation in adolescence (<18 years) or young adulthood and (3) knowing adolescent PWID in personal or injection networks. For this study, we defined adolescence as <18 years (using the legal definition of adulthood in India). Further, we defined young adulthood as the period between 18-24 years. In expanding our age criterion for recruitment, we used current and past iterations of “youth” in India[29]. We excluded participants < 18 years due to ethical considerations regarding informed consent.

Recruitment was performed face-to-face by facilitators at NGOs providing services to PWID via ICCs. While all male PWID were receiving OST, we intentionally recruited participants that varied in visit attendance levels (regular or irregular). Regular receipt of OST was defined as attendance >15 days each month. Given limited female-only programs, we recruited all female participants from a single site in Imphal, a few of whom were receiving OST. To explore staff perspectives at each NGO, staff present on the day of PWID FGDs participated in separate FGDs. Participants were provided a small monetary compensation for their time and transport needs. All PWID and providers who were approached accepted participation in the study.

The YR Gaitonde Centre for AIDS Research and Education and the Johns Hopkins Medical School institutional review boards provided study approval and ethical oversight.

### Patient and Public Involvement

Patients or the public were not involved in the design, or conduct, or reporting, or dissemination plans of our research.

### Data collection

We used a grounded theory approach[30] to explore three areas specific to adolescent and young adult PWID, with the goal of developing an explanatory model for engagement with services: (1) injection initiation experiences, (2) motivating factors and perceived barriers to receiving harm-reduction services, and (3) perspectives on service gaps and types of interventions needed.

Specific to harm-reduction services, we explored receipt of OST in greater depth than SSP for several reasons. First, Indian studies describe low OST participation among those who are opioid dependent (about 5%)[13,31]. Second, prior studies among PWID in India and elsewhere describe benefits of OST in ameliorating high-risk behaviors[16,32]. Third, NACO aims to expand OST participation among PWID[31,33]. We explored these areas in the context of the life course theory[34,35], recognizing late adolescence and young adulthood as stages wherein behaviors impacting later adult outcomes originate, presenting opportunities for intervention. Open-ended questions included in the FGDs were initially pilot tested in two study sites (Aizawl and Amritsar). Feedback was sought from PWID and providers who participated in the pilot FGDs regarding language, cultural sensitivity and pertinent questions to be additionally included in the final FGD interview guide.

Eights FGDs (5 PWID, 3 staff) were conducted between February 2018 and January 2019. Each PWID FGD comprised 8-11 participants and included younger and older PWID. Staff FGDs comprised 2-4 participants. In Imphal, FGDs were conducted separately for male and female PWID. Information regarding the study was provided ahead of the FGDs during recruitment of potential participants which was re-iterated again before FGDs were conducted. Participants were informed that their participation was voluntary and that decision either to participate or not would not impact their care. Participants were also informed of various safeguards to maintain their confidentiality. All participants provided verbal informed consent and were offered a small monetary incentive for their participation. FGDs were conducted in a private space within the NGO’s premises. With assistance from facilitators trained in qualitative research, FGDs were conducted by L.G (corresponding author) in the local dialect. L.G introduced herself as a pediatric infectious diseases physician seeking to develop services for young PWID in India. Each FGD took place until saturation of themes was achieved (this typically occurred over a span of 1-1.5 hours). FGDs were audio-recorded, transcribed, and translated to produce de-identified English-language transcripts.

FGD guides contained semi-structured questions to obtain narrative information in the three areas of interest. Questions regarding injection initiation elicited personal experiences and participants’ perceptions of what drives young people to initiate injection, as well as assessed injection network characteristics of adolescents (including how and where they obtain drugs and injecting equipment). Motivating factors and perceived barriers to receiving harm-reduction services were explored by asking about personal experiences in relation to peri-injection initiation (i.e., the time immediately before and after injection initiation) and after established injection behavior. Finally, participants were asked to identify gaps in services for young PWID and suggest services and interventions to increase engagement in relation to temporal stages of injection: pre-injection initiation, peri-injection initiation, and after established injection behavior.

### Data Analysis

Deductive and inductive approaches were used for thematic analysis. We created an initial coding scheme based on collective discussion within the research team and findings from similar studies. Two researchers (LG and CM) independently coded FGD transcripts, and met to identify new themes, reconcile discrepancies, and produce a final coding scheme (Supplementary Table 1). Transcripts were evaluated by a third researcher (AH) who arbitrated the code reconciliation process and ensured consistency in code application. Excerpts were randomly selected to calculate Cohen’s kappa statistic for inter-rater reliability of code application; this was found to be consistently ≥0.6.

Dedoose software (Sociocultural Research Consultants, LLC, Los Angeles, California) was used to generate descriptive statistics on the number of times each code appeared in a given FGD.

These descriptive statistics facilitated identification of common or salient themes and examination of relative differences in the prominence of specific themes by gender and across study sites.

We then conducted a deeper analysis with the goal of identifying interconnections between themes to develop a broader explanatory model for understanding engagement in services at each stage of injection. To do this, we re-evaluated common or salient themes from our initial coding to ascertain larger categories into which themes clustered.

### Data sharing

De-identified transcripts of FGDs are available by emailing the corresponding author.

**Supplementary Table 1.**
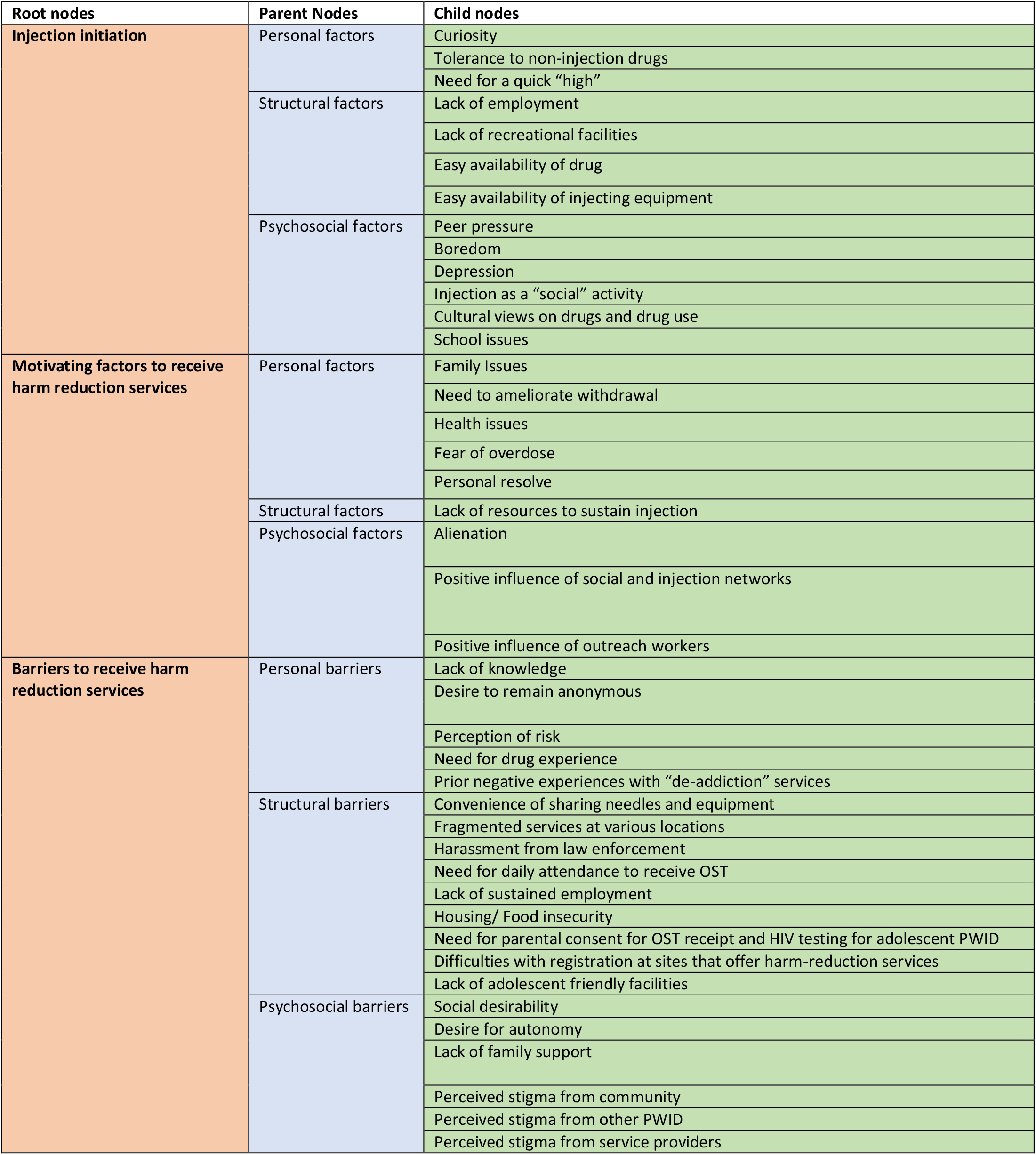
Coding scheme used to analyze focus group discussion transcripts

## RESULTS

### PARTICIPANT CHARACTERISTICS

A total of 43 PWID and 10 staff participated in FGDs (Table 1). The median age for male PWID was 27 years (range 20-30 years), while the median age for female PWID was 31 years (range 28-35 years). Half of PWID were receiving OST regularly. Staff participants included program managers, OST nurses, counselors and outreach staff.

**Table 1.** Descriptive characteristics of PWID participants

| Descriptive characteristic | N (%) (All sites) | Aizawl | Imphal | Amritsar |
| --- | --- | --- | --- | --- |
| Male | 35 (81%) | 11 | 8 | 16 |
| Female | 8 (19%) | -- | 8 | -- |
| Age |  |  |  |  |
| 18-24 years | 10 (23%) | 4 | 0 | 6 |
| 25-30 years | 28 (65%) | 7 | 11 | 10 |
| 31-35 years | 5 (12%) | -- | 5 | -- |
| Median age of injection initiation, Male PWID; (Range) | 20 years (12-24 years) | 18 years (12-23 years) | 18.5 years (16-22 years) | 22 years (16-24 years) |
| Median age of injection initiation, Female PWID; (Range) | 20.5 years (16-24 years) | -- | 20.5 years (16-24 years) | -- |
| Injection initiation in adolescence (<18 years) | 10 (23%) | 4 | 3 (Males)<br>2 (Females) | 1 |

## INJECTION INITIATION EXPERIENCES

Injection initiation experiences differed by gender and the time in life at which they were experienced. As such, we describe experiences unique to male PWID initiating injection in young adulthood, male PWID initiating injection in adolescence, and female PWID. Representative quotes are presented in Table 2.

**Table 2.** Factors leading to initiation of drug use and injection

| STRUCTURAL FACTORS | Representative quotation |
| --- | --- |
| Easy availability of drugs | **Q1.** Punjab is adjoining with Pakistan. Drugs are cheaper here than in other states and easily available. Drugs are like a fashion and easily available. Politicians and police indulge in the business of drugs -- Male PWID, Amritsar |
| Diversion of pharmaceutical drugs | **Q2.** The drug stores are selling (pharmaceutical drugs) to some bad medical representatives who then supply them to the peddlers – Male PWID, Aizawl<br>**Q3.** Pain tablets (from private de-addiction centers) are also easily available. Teenagers are injecting them after crushing –Male PWID, Amritsar |
| Unemployment | **Q4.** To be employed is a very big problem in our state because it is a very corrupt place. So, it's a main problem – Male PWID, Imphal<br>**Q5.** Some start injecting for fun, but some start injecting because they are unhappy and depressed due to unemployment, family problems, love failure – Male PWID, Amritsar |
| PERSONAL FACTORS |  |
| <sup>Y</sup>Curiosity and experimentation | **Q6.** Experimentation is there, excitement is there. Teenagers easily indulge because due to their growing age they are not able to recognize what is right or wrong for them. They easily adopt those things in which their peers are involved. When peer group forces them to use drugs, they can't refuse – Outreach staff, Amritsar |

| **PSYCHOSOCIAL FACTORS** |  |
| --- | --- |
| <sup>Y</sup>**Frustrations with actual lived realities** | **Q7.** *Because of globalization and media invasion they see what's going on around in big cities and want to follow the trend but the political and economic condition in Manipur is in contrast to what they expect, which is frustrating for them – Program manager, Imphal* |
| <sup>Y</sup>**School issues** | **Q8.** *Some start using after finishing school. In Punjab there is trend to go abroad after 12<sup>th</sup> grade. They do not want to do any work for the short-term. In that period, they easily indulge in drug addiction – Program manager, Amritsar*<br>**Q9.** *I started using drugs at the end of the year after I finished the school year. It was fun, as I got to hang out with my friends and got to talk about drugs. Then I dropped out after 3-4 years. I dropped out by myself because of the drugs – Male PWID, Aizawl* |
| <sup>Y</sup>**Desire to belong in peer groups** | **Q10.** *It depends on the group they associate with. When any sort of celebration occurs with friends who are already using drugs they can start taking too – Male PWID, Imphal* |
| <sup>Y</sup>**Notions of masculinity and normal behavior** | **Q11.** *Teenage boys who don't smoke weed or use other drugs think that it is not normal. They want to fit in – Male PWID, Aizawl* |
| <sup>W</sup>**Family issues (stigma of divorce and widowhood)** | **Q12.** *Mainly to escape from family problems. After getting divorced from my husband I was unhappy and had many problems, so I took drugs. – Female PWID, Imphal* |
| <sup>W</sup>**Differential power structures in relationships** | **Q13.** *I was used as a messenger to buy drugs for my husband and then later I got involved in the drug habit – Female PWID, Imphal*<br>**Q14.** *There are instances where partners come to purchase drugs together and at other times it's the boyfriends who send their girlfriends to purchase drugs. It's risky for males to purchase drugs while it's easier for girls – Female PWID, Imphal* |

<sup>Y</sup>Factors unique to adolescents; <sup>W</sup>Factors unique to female PWID

### Adult male PWID

Across all sites, structural factors were among the major drivers of injection initiation for adult male PWID. Easy availability of drugs was cited as a key reason for drug use (injection and non-injection) in all cities (Table 2, Q1). While participants purchased heroin primarily from peddlers, they described a local industry built on diversion of pharmaceutical drugs from pharmacies and private “de-addiction” centers despite prescription drug regulations (Q2-3).

> “*Medical store persons are getting business. They are there to earn money. Nobody monitors them on a regular basis. Most of the drugs are manufactured for good medical use but they (drug stores) are using them for different purposes. Ideal thing is that nobody can get medicine without doctor prescription but in India everybody is doctor of their own. Anyone can purchase drugs happily” – Program manager, Amritsar*

In all cities, unemployment served as a structural driver for drug use, by adversely affecting young peoples’ mental health. Along with family turmoil and relationship failures, unemployment led young people to use drugs as a “refuge”, due to the euphoric feelings they offered (Q4-5).

### Adolescent male PWID

In all cities, personal factors shaped injection initiation during adolescence. Participants described being inherently curious in adolescence, leading to experimentation with drugs (Q6). In the northeastern cities, staff reported that a disconnect between young peoples’ life expectations and their lived realities often led to a sense of frustration (Q7). Transitions (e.g., between school years) were periods of adolescent vulnerability (Q8-9). Participants identified poor school attendance and drop-out as increasing drug use risk in some individuals. In Aizawl, where all PWID had at least secondary school education and were in school during injection initiation, many noted that it was drug use that led them to drop-out (Q9).

Specific psychosocial factors unique to adolescents, such as the desire to belong in peer groups (Q10), as well as notions of what constituted masculinity and normal behavior (Q11), also contributed to injection initiation in all cities. Participants described adolescent injection networks as comprised of peers with slightly older members initiating younger users. Mobile phones and the internet fostered connectedness in networks. In contrast, participants described the interactions of adolescent PWID with much older established PWID (who they termed “hardcore”) as being purely transactional (i.e., occurring in the context of procuring drugs). Participants acknowledged a “code of conduct” that precluded seasoned PWID from teaching adolescents how to inject.

> *“Some teenagers use drugs because they want to feel confident to approach girls, confidence for exams. Some teenagers feel that being high is “cool” and some think that drugs help them feel manly” – Male PWID, Aizawl*

Participants in all cities described a rapid transition from non-injection to injection behavior due to the development of drug tolerance and the need for economic efficiency. Specific to the northeastern states, although participants noted that the easy availability of heroin facilitated initiation to injection directly (without a preceding period of non-injection drug use), participants indicated that the vast majority of young PWID in their networks had transitioned to injection after using pharmaceutical opioids and heroin via other routes. Participants described adolescents as initiating and continuing injection in locations where they would not be identified by family members (i.e., hotspots and peddler joints). Pharmacies were described as the primary sources of needles and syringes for male adolescents.

### Female PWID

Female PWID and staff described psychosocial factors as almost exclusively driving injection initiation among young women. While female participants described trajectories of adolescent experimentation that were similar to those described by male PWID, they also described stigma related to divorce and widowhood as leading to depression and subsequent drug use among older women (Q12).

Across all ages, drug use in women occurred in the context of differential power structures in relationships. Women related experiences of being introduced to drugs by their husbands or boyfriends, for whom they often procured drugs (Q13-14). PWID who were sex workers described a vicious cycle of sex work and drug use. Women depended upon informal sources, including partners, to obtain injecting equipment and drugs.

> *“My friend used me to buy and sell drugs as well as earn money for her as a sex worker. I was induced by my friend to use drugs so that I can be utilized as a tool for earning money for her”*
>
> *– Female PWID and sex worker, Imphal*

## MOTIVATING FACTORS AND PERCEIVED BARRIERS TO RECEIVING HARM-REDUCTION SERVICES

Motivating factors for receiving harm-reduction services did not vary by gender, therefore their findings are described together. In contrast, barriers to receiving and being retained in harm-reduction services varied considerably by gender and the time of life in which they were experienced, so we describe these barriers separately for adult male PWID, female PWID, and adolescent PWID. Related quotations are presented in Table 3, 4 and 5.

**Table 3.** Motivating factors to seek harm reduction services

| **PERSONAL FACTORS** | **Representative quotation** |
| --- | --- |
| **Health issues** | **Q15.** *My condition was very bad, health was poor, and family was not taking care of me. I was having abscesses on my arms. Then I came to know about OST and started it – Male PWID, Amritsar* |
| **Fear of death** | **Q16.** *I have taken drugs for a long time. One of my friends died due to overdose. I was afraid and I decided to take OST treatment – Male PWID, Amritsar* |
| **Fear of acquiring infections such as HIV** | **Q17.** *To reduce risk of HIV and any other infection, I decided to get clean needles and also OST – Male PWID, Amritsar* |
| **Withdrawal and dwindling money** | **Q18.** *I have found big relief now with OST. Earlier I was very unhappy due to withdrawals. Mostly I was having shortage of money and I was sharing drugs and equipment with my friends – Male PWID, Amritsar*<br>**Q19.** *Deteriorating health, no support from family and lack of money to afford heroin compelled me to choose OST – Female PWID, Imphal* |
| **Diminished withdrawal and urge to inject following OST** | **Q20.** *After taking OST the urge to use previous drugs have subsided, there is no physical pain and suffering, no withdrawal syndrome, health condition normalizes – Male PWID, Amritsar*<br>**Q21.** *It (OST) lasts longer – better than before when I used to take four or five hits of heroin in a day. Now I don't feel very dejected and my mind is fresh. I can spend time with family and friends – Female PWID, Imphal* |
| **PSYCHOSOCIAL FACTORS** |  |
| **Alienation from family and community** | **Q22.** *I was stealing money and other things from my own home and always fighting with my family. Every time I was thinking about drugs only – Male PWID, Aizawl* |
| **Desire for social acceptance** | **Q23.** *To escape the stigma associated with injecting heroin, and to at least feel accepted by society, plus declining money and family conditions compelled me to switch to Methadone – Female PWID, Imphal*<br>**Q24.** *I have been tagged as “Addict/ Amli”. Nobody wants to sit with me – Male PWID, Amritsar* |
| **Improved social functioning and financial independence** | **Q25.** *Since I started taking this medicine, people are more affectionate towards me, they welcome me and trust me. They even offer me jobs whenever possible. Earlier people did not trust me and did not welcome me. They were afraid I'll steal stuff from them. Now that I take OST, I can make my own future. Now my mother trusts me since I quit drugs and started working. She even believes it's because of taking this medicine that I'm able to work like a normal person. Now everyday she is concerned and reminds me to go and take OST – Male PWID, Imphal* |
| **Influence of peers in networks** | **Q26.** *Here my peer group is changed. My daily schedule is better now. The boys outside they have no schedule. They are only busy with their drugs – Male PWID, Amritsar* |
| **Influence of TI staff** | **Q27.** *They (staff) encourage us in very good manner and behave with us like family – Male PWID, Amritsar* |

**Table 4.** Barriers to seek harm-reduction services, specifically OST among male and female PWID

| **PERSONAL FACTORS** | **Representative quotation** |
| --- | --- |
| **Convenience of sharing needles** | **Q28.** *It is convenient to share needles which is why we don't get new needles – Male PWID, Aizawl* |
| **STRUCTURAL FACTORS** |  |
| **Lack of access to clean equipment during withdrawal** | **Q29.** *Peddlers they keep syringes and they give needles. Some inject and just get a fix somewhere hidden in the corner of the room and if the next person comes, because of the withdrawal they just quickly wash up with any available water and push. That is a more common practice here causing rapid hepatitis C to spread among injectors – Outreach worker, Imphal* |
| **Distance to OST centers** | **Q30.** *Major challenge is the distance to some OST centers. Especially since there are not enough of them. Some do not want to spend money daily on transport – Male PWID Amritsar* |
| **Need to be present daily to receive OST** | **Q31.** *My work starts at 8:30 AM. Sometime I face problem like if I am little late here then I should be absent from my work that day – Male PWID, Amritsar*<br>**Q32.** *Sometimes we have to go to meet my relatives who are in other cities, but we cannot stay there. There should be provision to get 2-3 days medicine for home, if we are going out of city – Male PWID, Amritsar* |
| **Delays in obtaining OST due to fragmented services** | **Q33.** *Main problem is that we have to wait for 3 days to start medicine. One day for doctor and on second day for testing, we need to go to the government hospital. The people who decide to leave drugs may have not an issue with waiting 3 days, but others may get frustrated and think there is no advantage in waiting. There is need to appoint full time doctor for induction – Male PWID, Amritsar* |
| **Police harassment** | **Q34.** *It's the police harassment when they catch me with the advance dose, I take home. They don't allow me to call the OST center to validate the medicine. Instead they will detain me and call my family and try to extort money. It's the situation here. Also, the medication sometimes has 'OST' label but many times it does not have label. Even when the medicines have labels, they will not listen to me and sometimes beat me – Male PWID, Imphal* |
| **PSYCHOSOCIAL FACTORS** |  |
| **Lack of family support** | **Q35.** *Initially some clients come regularly but when they face peer pressure among their community, they become irregular. Some of their family members do not support them. Some of them have family disputes. Whereas those who come regularly, they have a lot more family support – OST nurse, Amritsar* |
| **Stigma from family, community, healthcare workers towards male PWID** | **Q36.** *People who live in my locality, sometimes they used to mock me: "Where today you are going?" "Are you going to get some to the peddler with how much money?" "Do you need any money?" – Male PWID, Imphal*<br>**Q37.** *Programs implemented by NGOs comparatively have less stigma than government setup. Clients fear that if they go to government centers they might be labeled. Staff are less cooperative there – Program manager, Amritsar*<br>**Q38.** *They do not guide us properly. Behavior of staff (at government center) is rude and timing is very strict there – Male PWID, Amritsar* |
| **Stigma from family, community towards female PWID** | **Q39.** *Generally, a woman's reputation is always at question – and when she's a drug user, even after coming out of a rehabilitation center, the family and also the locality does not accept them – Female PWID, Imphal* |
| **Harassment from male PWID** | **Q40.** *It's inconvenient to take Methadone among the male crowd. We get harassed and teased, and sometimes we get followed thinking we'd be easy prey for sex – Female PWID, Imphal* |

**Table 5.**
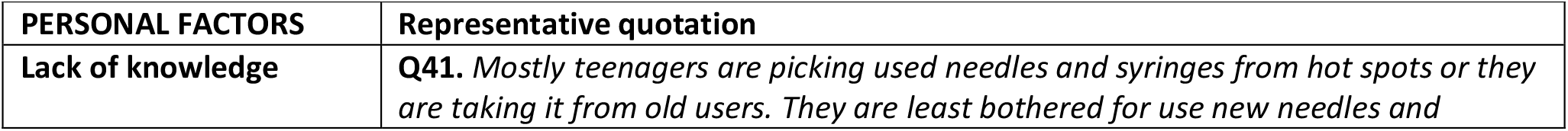
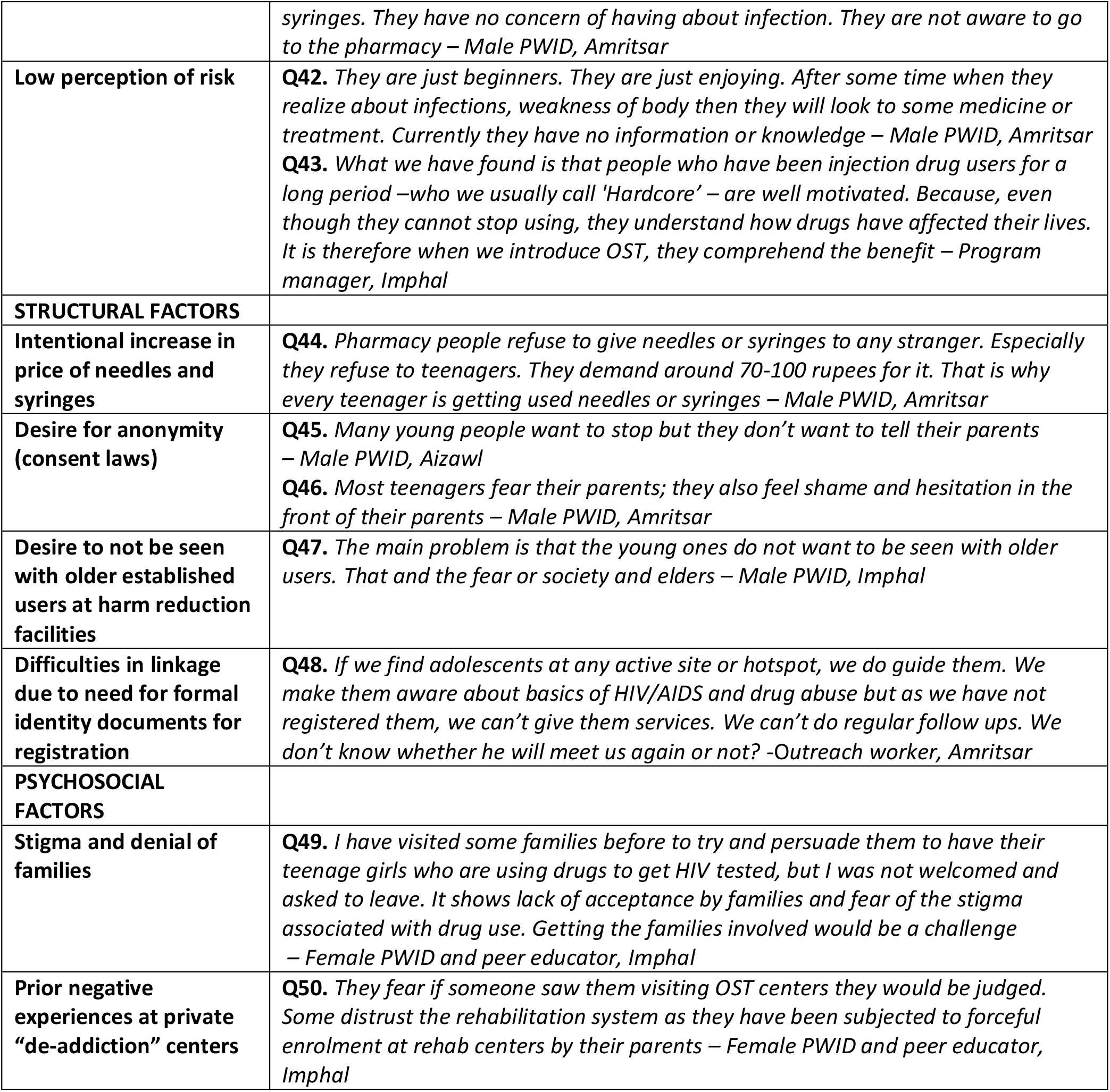
Barriers to seek harm-reduction services, specifically OST among adolescent PWID

| PERSONAL FACTORS | Representative quotation |
| --- | --- |
| Lack of knowledge | **Q41.** Mostly teenagers are picking used needles and syringes from hot spots or they are taking it from old users. They are least bothered for use new needles and |
|  | *syringes. They have no concern of having about infection. They are not aware to go to the pharmacy – Male PWID, Amritsar* |
| **Low perception of risk** | **Q42.** *They are just beginners. They are just enjoying. After some time when they realize about infections, weakness of body then they will look to some medicine or treatment. Currently they have no information or knowledge – Male PWID, Amritsar*<br>**Q43.** *What we have found is that people who have been injection drug users for a long period –who we usually call 'Hardcore' – are well motivated. Because, even though they cannot stop using, they understand how drugs have affected their lives. It is therefore when we introduce OST, they comprehend the benefit – Program manager, Imphal* |
| **STRUCTURAL FACTORS** |  |
| **Intentional increase in price of needles and syringes** | **Q44.** *Pharmacy people refuse to give needles or syringes to any stranger. Especially they refuse to teenagers. They demand around 70-100 rupees for it. That is why every teenager is getting used needles or syringes – Male PWID, Amritsar* |
| **Desire for anonymity (consent laws)** | **Q45.** *Many young people want to stop but they don't want to tell their parents – Male PWID, Aizawl*<br>**Q46.** *Most teenagers fear their parents; they also feel shame and hesitation in the front of their parents – Male PWID, Amritsar* |
| **Desire to not be seen with older established users at harm reduction facilities** | **Q47.** *The main problem is that the young ones do not want to be seen with older users. That and the fear or society and elders – Male PWID, Imphal* |
| **Difficulties in linkage due to need for formal identity documents for registration** | **Q48.** *If we find adolescents at any active site or hotspot, we do guide them. We make them aware about basics of HIV/AIDS and drug abuse but as we have not registered them, we can't give them services. We can't do regular follow ups. We don't know whether he will meet us again or not? -Outreach worker, Amritsar* |
| **PSYCHOSOCIAL FACTORS** |  |
| **Stigma and denial of families** | **Q49.** *I have visited some families before to try and persuade them to have their teenage girls who are using drugs to get HIV tested, but I was not welcomed and asked to leave. It shows lack of acceptance by families and fear of the stigma associated with drug use. Getting the families involved would be a challenge – Female PWID and peer educator, Imphal* |
| **Prior negative experiences at private “de-addiction” centers** | **Q50.** *They fear if someone saw them visiting OST centers they would be judged. Some distrust the rehabilitation system as they have been subjected to forceful enrolment at rehab centers by their parents – Female PWID and peer educator, Imphal* |

### Motivating factors

PWID across all sites described personal factors as major reasons for engaging in harm-reduction services, particularly OST. These included motivation stemming from experiencing injection-related health complications (Table 3, Q15), fear of death after witnessing overdose (Q16), and the fear of acquiring infections such as HIV (Q17). In many cases, it was an immediate need to ameliorate withdrawal symptoms (in the context of financial constraints) that pressed many to seek OST (Q18-19). Conversely, participants who received OST regularly described benefits such as diminished urge to inject, lack of withdrawal symptoms, and improved physical health (Q20-21).

Deteriorating personal relationships with family members was the single most frequently cited psychosocial reason to seek OST (Q22); the desire to be accepted by their families and communities was universal (Q23-24). Conversely, improved social acceptance and financial independence served as motivators to remain engaged (Q25). Male PWID specifically described encouragement of members in their networks as key reasons for seeking services. Regular OST attendance resulted in a transformation of male PWID peer groups, influencing positive behavior changes (Q26). Encouragement from staff was also cited as a reason to remain engaged with services (Q27).

> *“In the past there was lot of fight at home due to my drug behavior. I was coming home every day at 11 or 12 at night with high drug influence. Then I come to know about OST and decided to start it. Now my family has no complaints because I am not demanding money from them” – Male PWID, Amritsar*

### Barriers to services for male PWID

In all cities, the main reason described for not accessing SSP was the convenience of needle sharing (Table 4, Q28). For example, staff described lack of access to clean needles and syringes during periods of withdrawal as a key factor for PWID choosing to share needles rather than access SSP (Q29).

Both structural and psychosocial factors served as major barriers to receiving OST. With regard to structural barriers, there were considerable differences by region. In Amritsar, participants described physical distance as a barrier to reaching OST centers (Q30). Additionally, the need for daily attendance to receive OST was expressed by all participants as disruptive to social and work schedules (Q31-32), leading some to simultaneously inject while receiving OST. Others described delays and fragmented services. For example, some participants described not having a full-time physician leading to delays in OST induction, and to attrition of some PWID (Q33). In the Northeastern cities on the other hand, the unpredictable occurrence of civil unrest made it necessary for OST programs to offer take-home doses. PWID appreciated this flexibility but described harassment from law enforcement due to possession of OST medications (Q34).

> *“Sometimes I face problem like someday if I have urgent work and I need to go early but I have to come here daily. I can’t take home medicines. If I do not take medicine from here, then again, I need 1000 to 1500 rupees for drugs. Some boys are still using Injectables along with OST. Sometimes they feel lazy to come here” – Male PWID, Amritsar*

With regard to psychosocial barriers to receiving OST, staff observed that the biggest difference between those who remained engaged and those who did not was the presence or absence of family support (Q35). PWID across all sites generally agreed that lack of family support, and stigma from communities and healthcare workers were major challenges (Q36-38).

> *“Initially some clients come regularly but when they face peer pressure among their community, they become irregular. Some of their family members do not support them. Some of them have family disputes. Whereas those who come regularly, they have a lot more family support” – OST nurse, Amritsar*
>
> *“Family, means, about family I cannot say because still they believe that I’m the same person as I was in the past so that’s the problem. They do not know that I’m just changing after taking. They think what I am doing now is the same thing as the past. Sometimes I feel that it’s compulsory for family also just to come and see what we are doing here” – Male PWID, Imphal*

### Barriers to services for female PWID

PWID and staff described the multiple levels of stigma encountered by female PWID as being the biggest barrier to receive services. Female PWID encountered stigma not just for using drugs but also as divorcees, widows and/or sex workers (Q39). Female PWID reported receiving minimal family support, unlike some male PWID. As a result, they experienced housing and food insecurity. Females who accessed OST programs also described harassment from male PWID (Q40).

> *“They do not have any homes, no family support; most of the female injection drug users are homeless. If their families kick them out, society is not accepting of female injection drug users. They are the double stigmatized” – Program manager, Imphal*

### Barriers to services for adolescent PWID

With regard to personal factors, participants explained that adolescents lacked knowledge of and perceived need for harm-reduction services in the peri-initiation period (Table 5, Q41) because they did not envision themselves as being at-risk for negative consequences of drug use. Therefore, ongoing pleasurable experiences superseded their desire to seek harm-reduction services (Q42-43).

Participants described various structural barriers for adolescent PWID. While adolescents could buy needles and syringes from pharmacies, participants described instances where prices were intentionally increased, leading to needle sharing (Q44). Legal barriers for adolescents to receive services were implied through specific narratives. Consent laws in India stipulate that individuals <18 years of age require parental consent for HIV testing and OST. Participants at all sites described a stronger desire for anonymity among adolescent PWID when seeking services (Q45-46). Adolescents also did not wish to be seen with older “hardcore” users at facilities providing harm-reduction services (Q47). Staff described difficulties registering adolescents to access services due to a need for formal identity documents, which adolescents were unlikely to have (Q48).

> *“They won’t come because they don’t want to see you. They don’t want to meet you. They are afraid. They don’t want to be seen or known by people that they are doing drugs. Only by peddlers” – Male PWID, Aizawl*
>
> *“If we find adolescents at any active site or hotspot, we do guide them. We make them aware about basics of HIV/AIDS and drug abuse but as we have not registered them, we can’t give them services. We can’t do regular follow ups. We don’t know whether he will meet us again or not?*” −O*utreach worker, Amritsar*

With regard to psychosocial barriers, stigma and denial prevented family members of adolescent PWID from accompanying them to facilitate access to services (Q49). In addition, adolescent PWID who had previous negative experiences at private centers – that generally did not employ evidence-based treatments — were subsequently dissuaded from seeking harm-reduction services (Q50).

> *“They fear if someone saw them visiting OST centers they would be judged. Some distrust the rehabilitation system as they have been subjected to forceful enrolment at rehab centers by their parents” – Female PWID and peer educator, Imphal*

Female PWID noted that while older women had some degree of agency in seeking services for themselves, adolescent female PWID on the other hand had little agency even if they desired engaging in services.

> *“Some (adolescent females) would like to start OST. However, they do not want to go to OST centers. They ask us to bring back OST for them, or request that it be made available to them at home” – Female PWID and peer educator, Imphal*

### GAPS AND RECOMMENDATIONS FOR IMPROVING SERVICES

Participants identified gaps and offered recommendations for services, which are summarized in Table 6 and Figure 1. Gaps and recommendations varied by the stage that PWID were in their injection careers.

**Table 6.**
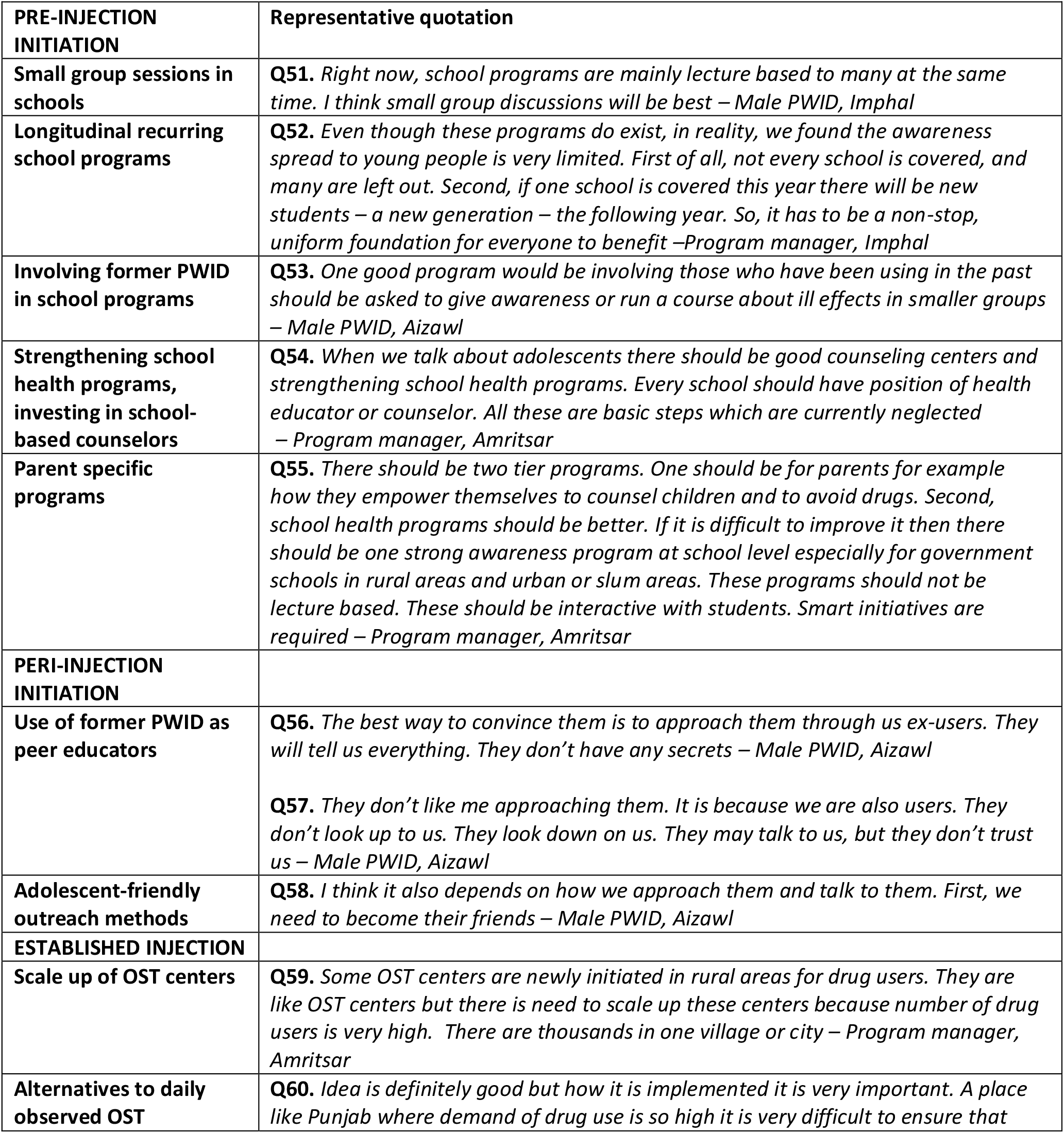
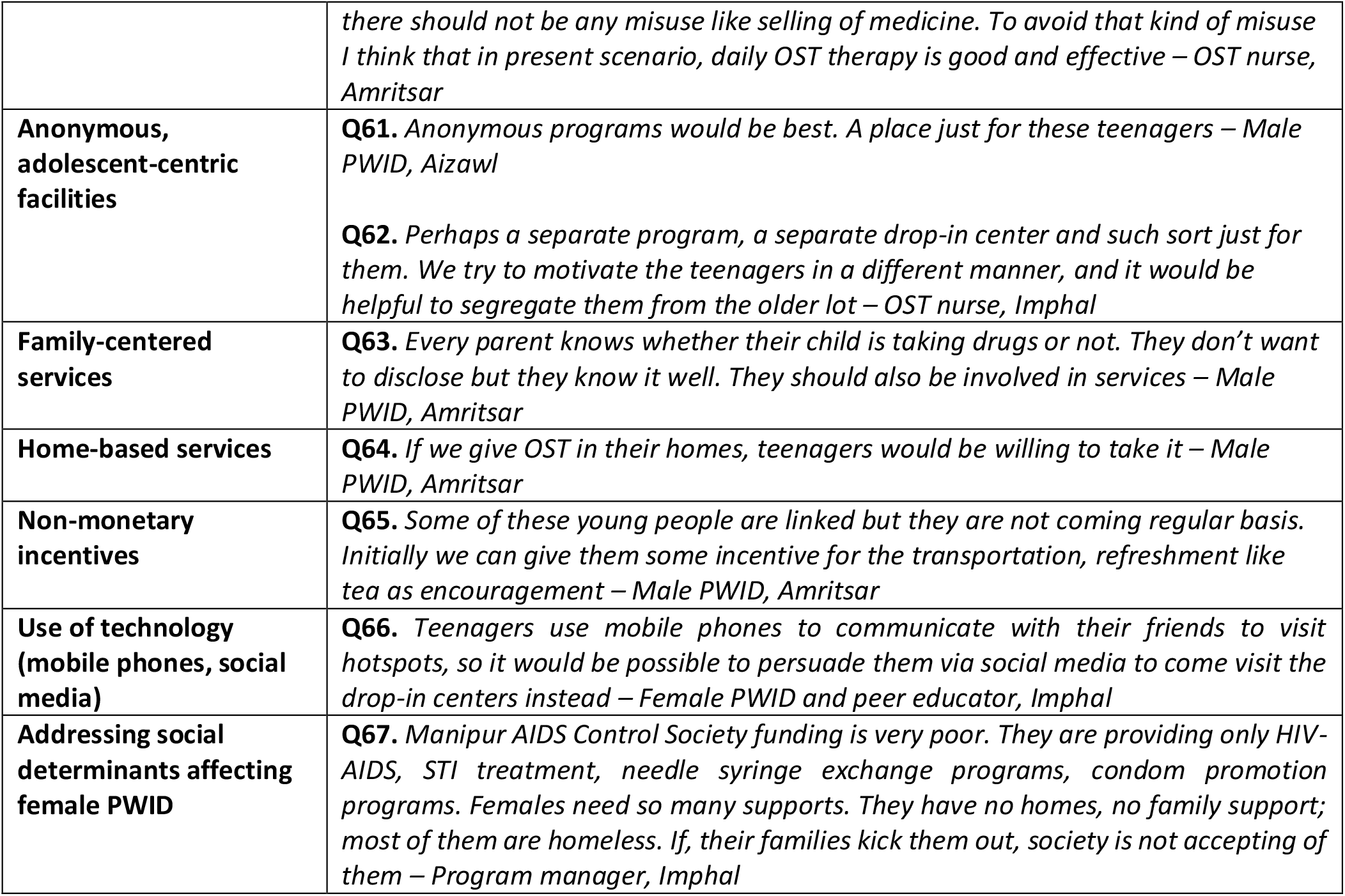
Recommendations for interventions and services

**Figure 1.**
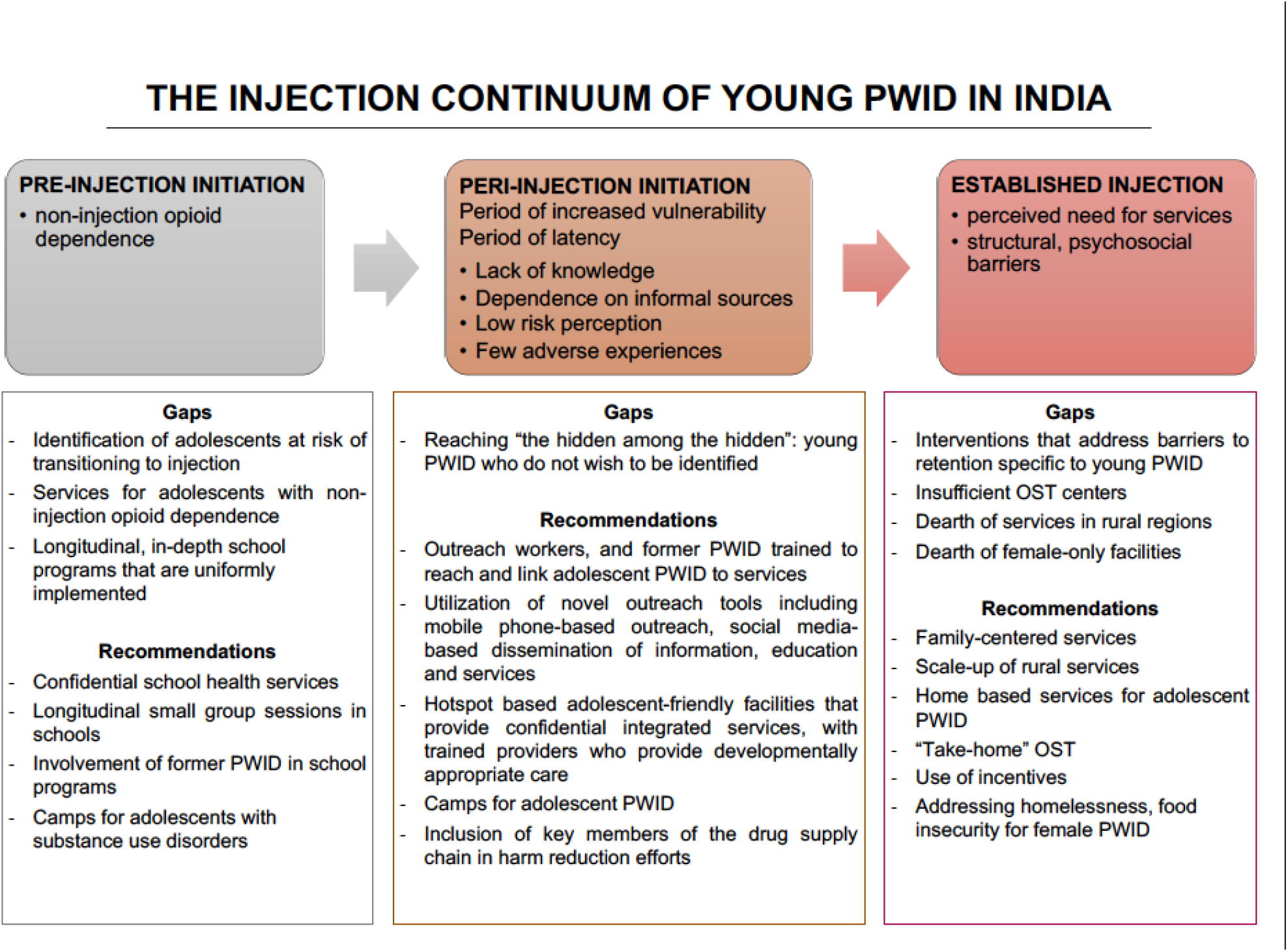
Explanatory model for engagement in services, service delivery gaps, and recommendations along the injection continuum. “Period of latency” refers to the period between injection initiation and established injection, where the is little perceived need for services.

### Pre-injection initiation

Gaps identified in the pre-injection initiation period included: (1) paucity of services for adolescents with non-injection substance use disorders, (2) failure to identify and support adolescents at risk of initiating injection, and (3) lack of uniformly implemented longitudinal substance-use education programs in high school. Participants suggested various ways to enhance school programs. These included conducting annual small group interactive educational sessions (Table 6, Q51-52), involving former PWID in education initiatives (Q53), investing in school-based counselors to address mental health issues that drive substance use in adolescents (Q54), and organizing camps for adolescents with substance use issues. In addition, programs for parents were viewed as being crucial to improve awareness and reduce stigma (Q55). Participants however noted that school-based programs would be of limited value to out-of-school youth at greatest risk for substance use.

### Peri-injection initiation

Male PWID across all sites described the peri-injection initiation period as a period of vulnerability, despite noting that young PWID generally do not experience significant complications of drug use during this time. Participants were pessimistic about engaging adolescents in this stage, highlighting the challenges of trying to engage those with limited insight.

Participants disagreed on the most effective interventions for this stage. Some participants saw value in ongoing interventions, such as training older former PWID to be peer-educators for adolescent PWID (Q56). Others felt that such interventions are unlikely to be effective, for a few reasons. Adolescent PWID were perceived to be reliant on peers – rather than older PWID— for injection initiation and social acceptance. Adolescents’ relationships with older users were also primarily transactional. Therefore, adolescent PWID were viewed as being inherently distrustful of older users (Q57). This in turn led to low motivation among peer-educators to engage adolescent PWID.

> *“Some of the peer educators teach them that how to inject safely and avoid abscesses. However, we don’t have enough time to teach them, talk with them. Some of them don’t take it seriously” – Male PWID, Amritsar*

Overall, participants recognized that engaging adolescent PWID could only happen after establishment of trust and suggested training peer-educators in adolescent friendly methods (Q58).

### Established injection

The main gap identified by participants at all sites was the lack of sufficient OST centers particularly in rural areas and the need for rapid scale-up given burgeoning injection in these areas (Q59). Specific to Amritsar, PWID highlighted the need to review daily observed OST therapy and implement “take home” dosing --a suggestion that was valuable to staff although many emphasized the need to also prevent misuse (Q60).

Some recommendations pertaining to adolescent PWID were applicable across the injection trajectory. At all sites, PWID identified the need for anonymous, adolescent-centric services, delivered ideally in locations close to hotspots (Q61-62). Although adolescents desired anonymity, participants emphasized that family-centered services by engaging parents collaboratively would sustain adolescent engagement (Q63). Some participants suggested alternatives to facility-based services, including exploring home-based service delivery models for adolescent PWID (Q64). Participants also recommended using non-monetary incentives (food, transport vouchers etc.) to sustain engagement (Q65). Additionally, harnessing technology (mobile phone and social media) to disseminate information about services was viewed to be powerful (Q66).

> “M*ostly they have smart phones. Most of them are using whatsapp, facebook etc. We need to use smart initiatives” –Outreach worker, Amritsar*

For female PWID, while the general lack of female-only facilities to provide harm-reduction services was recognized as a gap, participants noted that beyond provision of harm-reduction services, interventions need to address psychosocial determinants impacting outcomes among female PWID such as homelessness, food insecurity (Q67), and stigma and discrimination.

> *“Manipur AIDS Control Society funding is very poor. They are providing only HIV-AIDS, STI treatment, needle syringe exchange programs, condom promotion programs. Females need so many supports. They have no homes, no family support; most of them are homeless. If, their families kick them out, society is not accepting of them” – Program manager, Imphal*

### Explanatory model for engagement in services

In our explanatory model, we present key barriers to treatment engagement among young PWID in India, organized by stages within the injection continuum (figure 1). In the pre-initiation stage, the cross-cutting gap is the dearth of services and interventions aimed at preventing progression to injection, including the provision of services for non-injection opioid dependence. In the peri-injection initiation stage, lack of knowledge and dependence on informal sources of injecting equipment, combined with low risk-perception, result in low motivation to seek harm-reduction services. In the established injection stage, cumulative negative social and health consequences of IDU serve to motivate engagement with harm-reduction services, while structural and psychosocial barriers impede engagement. For adolescent PWID, lack of anonymous adolescent-friendly facilities impedes engagement across the continuum. Structural barriers (e.g., homelessness) and psychosocial barriers (e.g., stigma) reduce service engagement for all PWID, but disproportionately affect young female PWID.

## Discussion

In this study conducted in three cities in India, we provide insights into the diversity of injection experiences, facilitators and barriers to service engagement, and gaps in services faced by young PWID in India. These findings, along with direct recommendations for service delivery from PWID, inform a broader explanatory model for understanding harm-reduction service engagement by young PWID across the injection continuum.

To our knowledge, there are no prior Indian studies that examine the unique vulnerabilities of young PWID along the entire injection continuum. Addressing the needs of young people including young PWID, is critical to strengthen efforts to reduce HIV incidence[16]. Multiple recent large-scale trials aimed at increasing HIV testing or reducing HIV transmission via test-and-treat strategies have shown suboptimal results, partly due to poor engagement of hard-to-reach young individuals who have limited interactions with health services but are also drivers of transmission[36]. Our recently completed cluster-randomized trial evaluating the effectiveness of ICCs did not show an increase in community-level HIV testing among Indian PWID[17]. Post-hoc analyses revealed that the ICCs were reaching only a sub-set of the PWID in the community – for example, the median age of PWID registered at the ICCs was higher than the median age of PWID in the city suggesting that younger PWID may not be accessing these services. Incorporating strategies to ensure that these ICCs targeted all PWID sub-sets in the community may have improved HIV outcomes[37]. Similarly, failure to sufficiently engage young people has been cited as a potential reason for suboptimal reductions in HIV transmission in recent test-and-treat trials in sub-Saharan Africa[36,38–40].

Several motivating factors and barriers for young male PWID to receive services showed concordance with findings in other studies[41–44]. In the pre-injection initiation phase, the personal and psychosocial factors shaping progression from unaddressed non-injection drug dependence to IDU – including curiosity and desire to belong in peer groups – were similar across cities and consistent with findings from previous studies among young PWID in India and elsewhere[45–47]. The peri-injection initiation phase was identified as a period of increased vulnerability, particularly for adolescents, owing to their lack of knowledge, reliance on informal sources for injecting equipment, and low perceived risk. A period of latency ensued after injection initiation, characterized by lack of readiness among young PWID to engage with harm-reduction services and a perception that such services are not needed. Conversely, after established injection, additive negative physical and psychosocial experiences increased motivation to seek harm-reduction services. At this stage of readiness, personal factors such as improved physical functioning, and psychosocial factors, in particular family support and social acceptance, facilitated engagement. Psychosocial factors impeding engagement, such as stigma and discrimination, and structural factors, such as distance to services and lack of sufficient services, have also been reported elsewhere[41–44].

Historically, treatment of substance use disorders in India has relied on inpatient models of care (“de-addiction” centers), largely through poorly-regulated private facilities, some of which rely on abstinence-only approaches[48]. Additionally, daily attendance (even after induction) to receive OST in government and NGO-based programs has been the norm[49]. Prior negative experiences with “de-addiction” programs and the need for daily attendance, breeding reliance on factors such as transportation to receive OST were identified as barriers to engagement. The tendency for young PWID to seek clean needles and syringes from convenient sources such as pharmacies and informal sources such as friends was also a manifestation of the inconvenience of receiving harm reduction services at facility-based service delivery models. While some programs already offer “take-home” OST (i.e., 5-7 days’ supply), expansion to all public sector programs is expected to be rolled out soon as part of the National AIDS Control Program along with flexible timings for OST centers. Such initiatives likely need to be accompanied by education of members of law enforcement as well as providers given that participants in our study reported experiencing harassment from the former when provided with “take-home” OST. Additionally, the use of vouchers (such as transport vouchers as suggested by participants) and initiatives such as secondary needle exchange programs could also be of value.

Some interventions suggested by our study participants to engage adolescent PWID have been proposed in other youth consultations[41]. In our study, strengthening school-based services to prevent substance use initiation and addressing non-injection opioid dependence in adolescents were key recommendations in the pre-injection initiation phase. Participants held the view that education as well as identifying adolescents with non-injecting substance use dependence via school-based programs would delay injection initiation and improve their linkage to services. Although historically the availability of certain forms of heroin suitable for injecting (e.g., Heroin No. 4) in the northeastern states has facilitated initiation to injection directly among previously drug naïve young PWID, participants in our study noted that a significant proportion of young PWID in their networks including themselves had experienced a period of dependence to non-injection opioids suggesting this phase to be an important period for intervention. While studies that examine the effectiveness of school-based programs and services for substance use have been conducted in other settings and report variable benefit [50–54], longitudinal studies that examine the impact of school-based interventions on substance use among adolescents in India are needed. It should also be noted that participants in our study acknowledged that school-based programs were less likely to impact substance use in non-school going youth. While there are efforts to increase outpatient substance use services in India’s public sector[48,55], few adolescent – specific outpatient services exist although development of adolescent-centric service delivery models is currently under consideration. Based on recommendations from our participants, such services may be more readily accessed if they are hassle-free, anonymous and physically close to where adolescent PWID congregate but also separate from locations where services are delivered to older PWID. Under India’s National Adolescent Health Program, adolescent-friendly clinics that are designed to offer confidential services, including HIV testing and substance use services, have been established[56]. However, age restrictions to receive HIV testing and OST without parental consent endure, and recent evaluation of these clinics showed poor attendance by adolescents[57,58]. Therefore, creative service delivery platforms as suggested by our participants incorporating features such as home-based OST, incentives for engagement, use of technology to disseminate information, and revision of consent laws could lead to implementation of effective and accessible services for adolescents. Although anonymous services for adolescents were emphasized, there was also unanimous agreement on the need to develop family-centered service delivery models. Finally, our FGDs suggest that service delivery models for female PWID will need to extend beyond service provision and consider the psychosocial and structural factors that disproportionately affect women. Indeed, several prior studies among female PWID highlight the significant disparities that exist in this population[26,59–64]. The published literature to date however contains few if any narratives from adolescent female PWID. While this group was not represented directly in our study, narratives from older female PWID suggest that adolescent females possibly experience even greater stigma and possess lesser agency to seek services. The unique vulnerabilities and needs of this group are worthy of separate investigation and beyond the scope of this manuscript.

Our study findings challenge some previously described methods for increasing engagement among young PWID, including use of older network members[65–69]. Participants expressed ambivalence regarding how best to utilize network members in earlier stages of the injection continuum, citing low motivation among young PWID and unfavorable relationship dynamics between young and older PWID. Several studies show that adolescent PWID prefer to obtain needles and syringes from pharmacies even where SSP services exist[41,70,71], a finding that also emerged in our study. Although diversion of pharmaceutical drugs was described at all study sites, participants did not recommend empowering pharmacists to deliver harm-reduction education to young PWID, a strategy that has been explored in other countries[72– 74]. This finding could be related to PWID viewing their relationships with individuals involved in diversion of pharmaceutical drugs and sale of injecting equipment as purely transactional.

Our study has some limitations. While we sought to recruit young adult PWID exclusively, only a third of our participants fell into this age group. We also did not recruit any adolescent PWID due to ethical constraints. Although all PWID we recruited had initiated injection in adolescence or young adulthood, the preponderance of older PWID in our study could have led to respondent and recall bias. Therefore, the narratives elicited from our participants may not fully represent the needs of adolescents. Future studies examining engagement in services should include adolescent PWID participants, a current challenge in India given consent laws.

We also recruited participants who were already receiving services. Hence, their narratives may not be representative of PWID who have never engaged with harm-reduction services. This recruitment strategy may also be a reason why we were unable to recruit a greater number of young adult PWID and further emphasize the broader challenge of engagement among this group of PWID. The focus group discussions elicited a broad range of narratives, as well as common experiences among PWID. However, this format precluded exploring sensitive issues such as inter-personal violence including sexual violence experienced by young PWID and how these impact engagement in services. Participant narratives may also have been influenced by social desirability bias. While we observed common themes across cities in this study, our findings are not generalizable to other cities in India.

Limitations notwithstanding, these findings represent some of the first qualitative data to explore engagement with services, specifically among young PWID in India. Our study also offers insights to support the development of adolescent-specific interventions and services along the injection continuum. The inclusion of multiple cities adds to the strength of our observations. Given high HIV incidence among young PWID in India, there is an urgent need to address their engagement with services in a specific, systematic and sustained manner. Services designed for young PWID need to be multi-pronged and implemented simultaneously across the injection continuum, addressing challenges in each phase. Scale-up of innovative, integrated and holistic service delivery models separate from those targeting older adults will be critical to accelerate reductions in HIV transmission and achieve UNAIDS’ 95-95-95 targets in India.

## Supporting information

COREQ checklist

## Data Availability

De-identified transcripts of FGDs are available by emailing the corresponding author.

## Acknowledgements

This project was supported by the National Institute on Drug Abuse of the National Institutes of Health (R01DA032059, R01DA041034, DP2DA040244 and K24DA035684). The project was also supported by a Harvard HIV T32 post-doctoral fellowship (T32AI007433) from the National Institute of Allergy and Infectious Diseases, the Harvard University Center for AIDS Research (CFAR), an NIH funded program (P30AI060354), The Johns Hopkins University CFAR (P30AI094189), The Elton John AIDS Foundation, The Aerosmith Research Endowment Fund and a Career Development Fellowship from Boston Children’s Hospital. Its contents are solely the responsibility of the authors and do not necessarily represent the official views of the National Institutes of Health. We thank the National AIDS Control Organization (NACO), India, all of our partner non-governmental organizations throughout India, and the participants, without whom this research would not have been possible.

## Author contributions

Lakshmi Ganapathi, Sion Kim Harris and Sunil Suhas Solomon conceived the study with support from Vinita Verma, Shobini Rajan, Gregory M Lucas, Shruti H Mehta, Kenneth H Mayer, Conall O’Cleirigh; Aylur K Srikrishnan coordinated participant recruitment; Lakshmi Ganapathi participated in the conduct of FGDs, analyzed the data and drafted the manuscript; Clarissa Martinez and Areej Hassan analyzed the data. Gregory M Lucas, Shruti H Mehta, Vinita Verma, Allison McFall, Kenneth H Mayer, Conall O’Cleirigh, Areej Hassan, Shobini Rajan, Sion Kim Harris and Sunil S Solomon provided key edits to the manuscript.

