## Supplementary material for "Young and invisible: an explanatory model for service engagement by people who inject drugs in India": COREQ checklist

### Consolidated criteria for reporting qualitative studies (COREQ): 32-item checklist

Developed from:
Tong A, Sainsbury P, Craig J. Consolidated criteria for reporting qualitative research (COREQ): a 32-item checklist for interviews and focus groups. *International Journal for Quality in Health Care*. 2007. Volume 19, Number 6: pp. 349 – 357

| No. Item | Guide questions/description | Reported on Page # |
|---|---|---|
| **Domain 1: Research team and reflexivity** | | |
| *Personal Characteristics* | | |
| 1. Inter viewer/facilitator | Which author/s conducted the inter view or focus group? | Methods, data collection, page 7 |
| 2. Credentials | What were the researcher's credentials? E.g. PhD, MD | Methods, data collection, page 7 |
| 3. Occupation | What was their occupation at the time of the study? | Methods, data collection, page 7 |
| 4. Gender | Was the researcher male or female? | Methods, data collection, page 7 |
| 5. Experience and training | What experience or training did the researcher have? | Methods, data collection, page 7 |
| *Relationship with participants* | | |
| 6. Relationship established | Was a relationship established prior to study commencement? | N/A |
| 7. Participant knowledge of the interviewer | What did the participants know about the researcher? e.g. personal goals, reasons for doing the research | Methods, data collection, page 8 |
| 8. Interviewer characteristics | What characteristics were reported about the inter viewer/facilitator? e.g. Bias, assumptions, reasons and interests in the research topic | Methods, data collection, page 8 |
| **Domain 2: study design** | | |
| *Theoretical framework* | | |
| 9. Methodological orientation and Theory | What methodological orientation was stated to underpin the study? e.g. grounded theory, discourse analysis, ethnography, phenomenology, content analysis | Methods, data analysis, page 8 |
| *Participant selection* | | |
| 10. Sampling | How were participants selected? e.g. purposive, convenience, consecutive, snowball | Methods, study population, page 6 |
| 11. Method of approach | How were participants approached? e.g. face-to-face, telephone, mail, email | Methods, study population, page 6 |
| 12. Sample size | How many participants were in the study? | Results, page 11 |
| 13. Non-participation | How many people refused to participate or dropped out? Reasons? | Methods, study population, page 6 |

| | | |
|---|---|---|
| *Setting* | | |
| 14. Setting of data collection | Where was the data collected? e.g. home, clinic, workplace | Methods, data collection, page 7 |
| 15. Presence of non-participants | Was anyone else present besides the participants and researchers? | Results, page 11 |
| 16. Description of sample | What are the important characteristics of the sample? e.g. demographic data, date | Results, page 11 |
| *Data collection* | | |
| 17. Interview guide | Were questions, prompts, guides provided by the authors? Was it pilot tested? | Methods, data collection, page 7 |
| 18. Repeat interviews | Were repeat inter views carried out? If yes, how many? | N/A |
| 19. Audio/visual recording | Did the research use audio or visual recording to collect the data? | Methods, data collection, page 7 |
| 20. Field notes | Were field notes made during and/or after the inter view or focus group? | N/A |
| 21. Duration | What was the duration of the inter views or focus group? | Methods, data collection, page 7 |
| 22. Data saturation | Was data saturation discussed? | Methods, data collection, page 7 |
| 23. Transcripts returned | Were transcripts returned to participants for comment and/or correction? | N/A |
| **Domain 3: analysis and findings** | | |
| *Data analysis* | | |
| 24. Number of data coders | How many data coders coded the data? | Methods, data analysis, page 8,9 |
| 25. Description of the coding tree | Did authors provide a description of the coding tree? | Methods, data analysis, page 8,9 |
| 26. Derivation of themes | Were themes identified in advance or derived from the data? | Methods, data analysis, page 8,9 |
| 27. Software | What software, if applicable, was used to manage the data? | Dedoose, page 8 |
| 28. Participant checking | Did participants provide feedback on the findings? | N/A |
| *Reporting* | | |
| 29. Quotations presented | Were participant quotations presented to illustrate the themes/findings? Was each quotation identified? e.g. participant number | Results, pages 11-24 |
| 30. Data and findings consistent | Was there consistency between the data presented and the findings? | Relationship to existing knowledge, page 27 |
| 31. Clarity of major themes | Were major themes clearly presented in the findings? | Results, pages 11-24 |
| 32. Clarity of minor themes | Is there a description of diverse cases or discussion of minor themes? | Results, pages 11-24; Discussion, pages 27-31 |
